# Scaled *m*-BonacciNet: A Recurrence-Guided Lightweight CNN for Efficient Medical Image Classification

**DOI:** 10.64898/2026.10.18.26362018

**Authors:** Gousia Habib, Shivani Thakur, Muhammad Shaban, Rehan Ahmad, Ishfaq Ahmad Malik

## Abstract

The *m*-Bonacci weighted sequence space 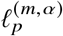 is introduced for *m* ≥ 2, 1 ≤ *p* < ∞, and *α* > 0. It is proved to be a Banach space isometrically isomorphic to *l*_*p*_, with an unconditional Schauder basis. The space is also reflexive and uniformly convex. Its strict inclusion chain, continuous dual, and asymptotic truncation estimate of order 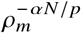 are established. Based on this framework, Scaled *m*-BonacciNet is proposed as a lightweight CNN that rescales Fibonacci-Net channels using the dominant *m*-Bonacci root and employs depthwise separable convolutions. Tribonacci-Net achieved 96.09% accuracy and 0.9581 macro F1-score on the 15-class Brain MRI dataset using 0.85 million parameters and 0.366 GFLOPs. Pentanacci-Net attained 95.87% accuracy on the 44-class dataset, reducing parameters and FLOPs by 47.10% and 44.48%. Hexanacci-Net achieved 83.75% accuracy and 0.7941 macro F1-score on CKD ultrasound.

## 1 Introduction

The significance of medical imaging lies in its contributions to illness screening, diagnosis, treatment strategy, and clinical assessment. Nevertheless, the escalating amount and complexity of imaging data can lead to time-intensive manual interpretation and vulnerability to observer variability. As a result, deep learning, notably thru convolutional neural networks (CNNs), has become a significant technique for medical image classification, detection, and segmentation, owing to its capacity to learn hierarchical representations from imaging data [1, 2]. However, the real-world implementation of CNNs in healthcare demands not just exceptional classification performance but also computational economy, reproducibility, and interoperability with resource-restricted clinical systems.

The increased architectural depth, channel width, parameter count, memory usage, and computational cost are often associated with the performance gains made by contemporary CNNs. Deployment on mobile devices, edge platforms, and clinical systems with constrained hardware resources may be restricted by these constraints. As a result, lightweight designs have been created to enhance the trade-off between accuracy and efficiency. ShuffleNet uses pointwise group convolution and channel shuffling [3], MobileNet uses depthwise-separable convolutions and width multipliers [4], and EfficientNet jointly scales network depth, width, and input resolution [5]. To lower processing and storage needs, model-compression techniques like pruning, quantisation, and weight sharing have also been applied [6]. However, manually created schedules, empirical scaling coefficients, or architecture-search techniques are typically used to determine the layer-wise channel configurations of these models.

Fibonacci-Net proposed a recurrence-guided alternative, where the number of convolutional filters is allocated following the Fibonacci cycle [7]. The number of filters is equivalent to the layer-wise channel allocation or network width, because each convolutional filter outputs a single output feature channel.

Such Fibonacci-based filter allocation offers an interpretable alternative to the standard channel-growth schedules, but is constrained to a set second-order recurrence and offers little flexibility to adjust the network capacity. Moreover, higher-order recurrence sequences tend to expand fast and could enlarge the model complexity if directly employed as filter counts. This leads us to the main research question of this study: ***Can a scaled*** *m****-Bonacci recurrence offer a controllable filter-allocation approach to reduce the CNN complexity while maintaining classification accuracy?*** We answer this question by introducing *Scaled m-BonacciNet*, a recurrence-guided lightweight CNN where the recurrence order *m* governs an order-dependent scaling coefficient via the dominant *m*-Bonacci root.

Let (*F*_1_, *F*_2_, … , *F*_7_) = (21, 34, 55, 89, 144, 233, 377) represent the original configuration of the Fibonacci-Net channel.

For *m* ≥ 2, the scaling coefficient is defined as 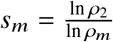.

where *ρ*_*m*_ is the dominant positive root of the characteristic polynomial associated with the *m*-Bonacci recurrence. The output-channel count of the *l*th convolutional block is then defined as

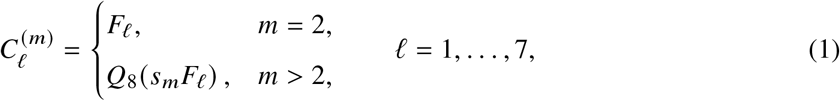

where *Q*_8_(·) rounds the scaled value to a valid multiple of eight. Therefore, the raw higher-order m-Bonacci sequence terms are not directly used as filter counts. The scaling is instead determined by the dominant root on the initial Fibonacci-Net channel setup. The resulting order-dependent channel allocation is paired with depthwise-separable convolution in the last convolutional blocks to reduce the model complexity. The main contribution of this work is a dominant-root-scaled channel-allocation framework for recurrence-guided lightweight CNN design. The suggested framework is an extension of Fibonacci-Net, where an explicit channel-scaling coefficient is derived by applying the recurrence order, and a common network topology is retained for the evaluated model versions.

Furthermore, the Banach-space structure, basis properties, stringent inclusion relations, continuous dual, and asymptotic truncation behaviour of the corresponding *m*-Bonacci weighted sequence space are examined. The proposed CNN versions are assessed in terms of classification performance, parameters count, MACs, FLOPs, model size and inference efficiency.

## 2 Background and Related Work

Recent progress in lightweight deep learning has been driven by advancements in mathematical modelling, efficient architecture design, and application-specific categorisation approaches. In this work, a (m)-Bonacci recurrence bridges these research directions to offer the mathematical basis of the proposed sequence space and a systematic rule of layer-wise channel allocation in a lightweight CNN. The present section focuses on three closely connected areas: sequence spaces based on recurrence and neural operators, lightweight CNNs with focus on recurrence-guided channel allocation, and efficient deep-learning methods for medical-image categorisation.

### Recurrence-Based Sequence Spaces and Neural Operators

The construction of Banach sequence spaces from matrices generated by special integer sequences has become an active topic in functional analysis. Recent studies have particularly focused on Fibonacci-, *q*-Fibonacci-, and Tribonacci-based matrix domains. Yaying et al. [8] introduced *q*-Fibonacci sequence spaces and investigated their Schauder bases, Köthe–Toeplitz duals, matrix mappings, and compact operators. Gökçe [9] developed absolute Tribonacci series spaces and established their BK-space structure, duals, summability properties, and compact and matrix operator characterisations. Similarly, Perdahlı et al. [10] studied Fibonacci–Euler sequence spaces and derived their topological properties, Schauder bases, duals, and associated matrix classes.

Earlier contributions established several important foundations for this line of research. Yaying et al. [11] constructed sequence spaces using a generalized Fibonacci difference operator formed from a Fibonacci band matrix and a triple-band matrix, and examined their topological properties, Schauder bases, Köthe–Toeplitz duals, matrix mappings, and compact operators. Yaying and Hazarika [12] introduced sequence spaces generated by a regular Tribonacci matrix and studied their inclusion relations, bases, duals, matrix transformations, and compact operators. Alp and Kara [13] further defined Banach sequence spaces through the domain of a Riesz–Fibonacci matrix and characterised their duals and corresponding matrix classes. Related work includes the study of absolute Fibonacci series spaces and their matrix and compact operators by Gökçe and Sarıgöl [14], as well as the domain of the Fibonacci band matrix investigated by Yaşar [15]. These studies demonstrate that recurrence-generated matrices provide a systematic framework for constructing new BK-spaces and analysing their topological, dual, summability, and operator-theoretic properties.

Recently, Yaying et al. [16] characterised matrix mappings and compact operators on Riordan-number sequence spaces. Simultaneously, Feischl et al. [17] related Banach fixed-point iterations to neural-operator approximation and derived convergence and error estimates. Current research however mainly treat single recurrence sequences independently and do not explore the effect of a changing recurrence order on the architecture of the generated spaces and the truncation behaviour of related neural operators.

### Lightweight CNNs and Recurrence-Guided Channel Allocation

The design of lightweight CNNs is a trade off between prediction performance, computational complexity, memory usage and inference time. Recent lightweight CNNs accomplish efficiency by tiny blocks, efficient convolutions and hardware-aware design. MobileOne [18] adopted structural reparameterisation to achieve low-latency inference. FasterNet [19] proposed partial convolution to reduce the duplicate computation and memory access. MobileNetV4 [20] used efficient bottleneck blocks, mobile attention, and hardware-aware architecture search for accuracy and latency trade-offs. More recently, LSNet [21] combines the large-field perception and small-kernel aggregation, and ShiftwiseConv [22] exploits sparse multi-path dependencies to efficiently achieve large-kernel effects. The practical efficiency is not only associated with computational complexity but also with memory access and inference delay, as shown by Korkmaz [23].

However, current architectures usually choose layer-wise channel widths manually, empirically or by architectural search. In contrast, the suggested method uses a scaled *m*-Bonacci recurrence, where *m* sets the channel growth pattern and *s* regulates the total network width. Thus it delivers an interpretable, math-ematically specified channel-allocation technique without any post-training compression or architecture search.

### Efficient Deep Learning for Medical-Image Classification

High performance classification with manageable computational and memory requirements is needed when deploying deep learning models in clinical settings. Naeem et al. [24] introduced a lightweight five-layer CNN for brain tumour detection with minimal MRI data. Appasami and Savarimuthu [25] optimised kernel, pooling and batch configurations to build a parameter efficient classifier for MRI. Dutta et al. [26] proposed ARM-Net, which combines attention, residual connections and multiscale feature extraction. Zahoor et al. [27] introduced a residual and regional CNN to capture tumour boundaries and textures. CerevianNet [28] employed a compact custom CNN with progressively increasing filters for multiclass brain tumour classification. Pimpalkar et al. [29] compared fine-tuned CNN and transfer-learning models for classifying normal kidneys, cysts, stones and tumours for kidney CT images.

While these methods yield good classification performance, they are mostly based on task-specific architectures, predefined filter schedules, attention modules or pretrained models. Conversely, we propose a unified and controllable filter-allocation technique via a scaled *m*-Bonacci recurrence. Its performance is assessed in terms of accuracy, macro F1-score, number of parameters, MACs, FLOPs, model size and inference efficiency on the brain MRI and kidney CT datasets.

Another closer approach is Fibonacci-Net [7] which allocates filter counts per layer in a Fibonacci sequence and employs depthwise-separable convolutions to lower computational cost. Tho approach introduces an interpretable recurrence-guided allocation strategy, but it is restricted to a fixed second-order recurrence and allows little control over the overall network breadth. The current study generalises this notion with a scaled *m*-Bonacci setting, in which the recurrence order *m* influences the filter-growth pattern and the scaling coefficient *s* regulates the overall width of the network. Thus, the suggested method provides a larger and more controllable family of the recurrence-guided lightweight CNNs.

## 3 Mathematical Formulation

The proposed framework contains two related components. First, an *m*-Bonacci weighted sequence space is introduced to characterize the growth and approximation properties associated with higher-order recurrences. Second, the dominant root of the *m*-Bonacci recurrence is used to construct an order-dependent scaling coefficient for rescaling the original Fibonacci-Net channel configuration. The resulting channel schedule is combined with depthwise-separable convolution to obtain a lightweight CNN architecture.

### 3.1 The *m*-Bonacci Sequence

For an integer *m* ≥ 2, the *m*-Bonacci sequence 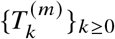 is defined by

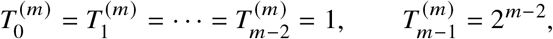

together with the recurrence

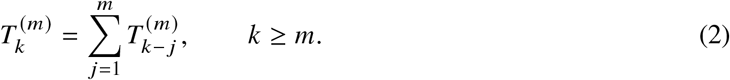

For *m* = 2, 3, and 4, these initial conditions generate the Fibonacci-, Tribonacci-, and Tetranacci-type sequences, respectively:

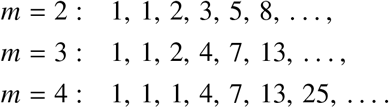

The characteristic polynomial associated with Equation (2) is *P*_*m*_(*x*) = *x*^*m*^ − *x*^*m*−1^ − *x*^*m*−2^ − … − *x* − 1. Let *ρ*_*m*_ > 1 denote the dominant positive root of *P*_*m*_. Then the sequence has the asymptotic form

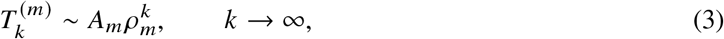

where *A*_*m*_ > 0 depends on the recurrence order and the chosen initial conditions. The dominant roots satisfy

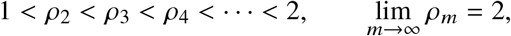

with *ρ*_2_ ≈ 1.618034, *ρ*_3_ ≈ 1.839287, *ρ*_4_ ≈ 1.927562, *ρ*_5_ ≈ 1.965948, *ρ*_6_ ≈ 1.983583. Thus, the asymp-totic growth rate of 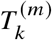 increases with *m* and approaches the limiting growth factor 2.

### 3.2 The *m*-Bonacci Weighted Sequence Space

Let *ω* denote the vector space of all real or complex sequences. For *m* ≥ 2, 1 ≤ *p* < ∞, and *α* > 0, we define the weighted *m*-Bonacci sequence space by

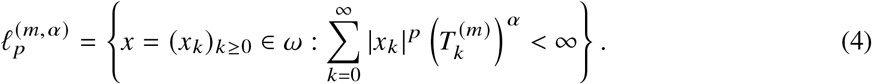

Equipped with the norm

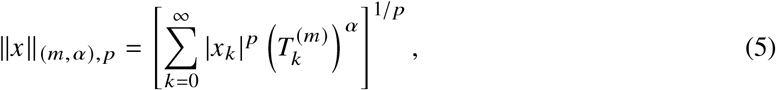

this space consists of all sequences whose *p*-th powers are summable with respect to the *m*-Bonacci weight

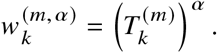

Thus, the parameter *m* determines the asymptotic growth rate of the weights, *α* controls the intensity of the weighting, and *p* specifies the underlying summability exponent.

### 3.3 Banach-Space Structure

#### Theorem 1.

*For m* ≥ 2, 1 ≤ *p* < ∞, *and α* > 0, *the space* 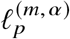 *is a Banach coordinate space and isometrically isomorphic to l*_*p*_.

*Proof*. The space (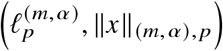, ∥*x*∥_(*m,α*), *p*_) is a Banach space is obvious. We show its isometrically isomorphic to *l*_*p*_. Define the linear mapping

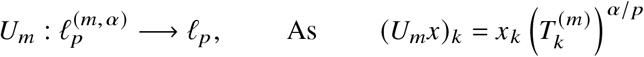

For 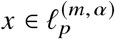,

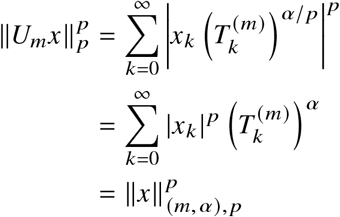

Hence, ∥*U*_*m*_*x*∥ _*p*_ = ∥*x*∥_(*m,α*), *p*_. Conversely, for *y* = (*y*_*k*_) ∈ *l*_*p*_, define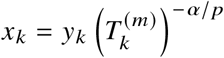.

Then 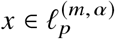 and *U*_*m*_*x* = *y*; therefore, *U*_*m*_ is a surjective linear isometry. Since *l*_*p*_ is complete, 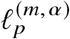 is complete as well. Moreover,

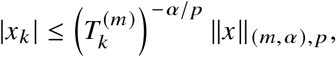

so every coordinate projection is continuous. Thus, 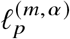 is a Banach coordinate space isometrically isomorphic to *l*_*p*_.

### 3.4 Strict Inclusion Across Recurrence Orders

#### Theorem 2.

*For m* ≥ 2, 1 ≤ *p* < ∞, *and α* > 0,

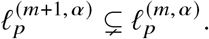

*Consequently*,

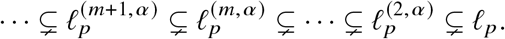

*Proof*. By Equation (3),

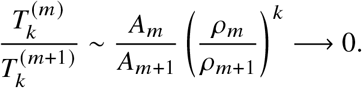

Therefore, there exists *D*_*m*_ > 0 such that 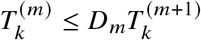 for all *k* ≥ 0. Thus, for 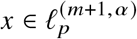,

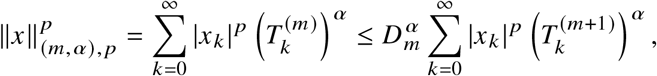

which proves the continuous inclusion. To establish strictness, define

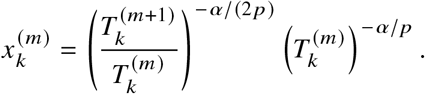

Then

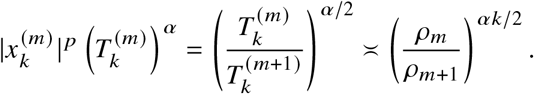

Since *ρ*_*m*_/*ρ*_*m*+1_ < 1, the corresponding series converges, and hence 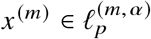. However,

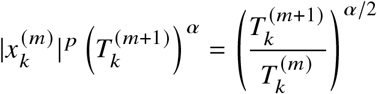

whose terms do not tend to zero.

Consequently, 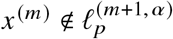. Therefore, the inclusion in Equation (2) is strict.

### 3.5 Basis and Geometric Properties

#### Theorem 3.

*The canonical unit vectors* {*e*^(*k*)^}_*k*≥ 0_ *form an unconditional Schauder basis for* 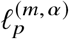 , *with unconditionality constant* 1.

*Proof*. For 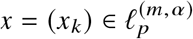 , define 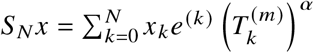.

Then,

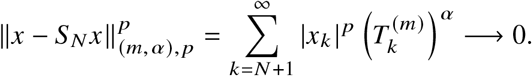

For every finite *E* ⊂ ℕ _0_ and every choice |*ε*_*k*_ | = 1,

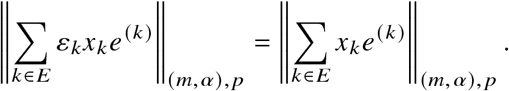

Thus, the basis is unconditional with constant 1.

#### Corollary 1.

*For* 1 < *p* < ∞, *the space* 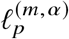 *is separable, reflexive, and uniformly convex*.

### 3.6 Continuous Dual

#### Theorem 4.

*Let p*^′^ *denote the conjugate exponent of p, that is* 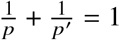. *Then, for* 1 < *p* < ∞,

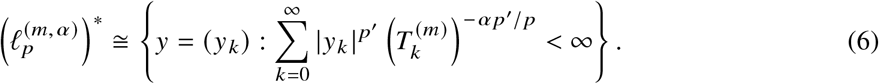

*Proof*. For *y* = (*y*_*k*_), define 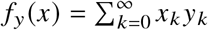. By H*ö*lder’s inequality,

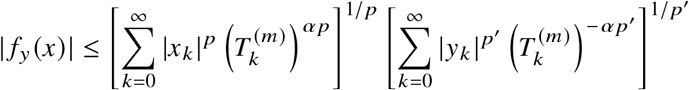

Therefore, every sequence satisfying the condition in Equation (6) defines a continuous linear functional on 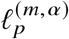.

Conversely, given 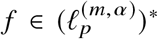, transfer *f* to *l*_*p*_ through the isometry *U*_*m*_. Since 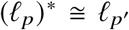 , the representing sequence belongs precisely to the weighted class specified in Equation (6). Hence the stated dual identification follows.

### 3.7 Truncation Estimate

The weighted-space structure yields an explicit truncation estimate in the unweighted *l*_*p*_-norm. Let

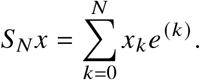

#### Theorem 5.

*For every* 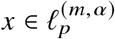 ,

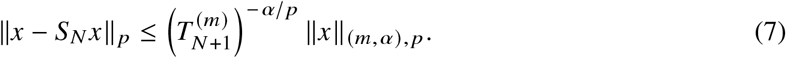

*Proof*. Since 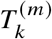 is non-decreasing,

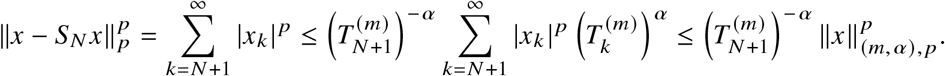

Taking the *p*th root proves Equation (7).

Using 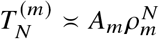 , there exists a constant *B*_*m,α, p*_ > 0, independent of *x* and *N*, such that

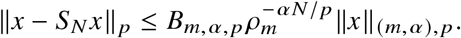

Consequently, a sufficient truncation dimension for ∥*x* − *S*_*N*_ *x*∥ _*p*_ ≤ *ε* is

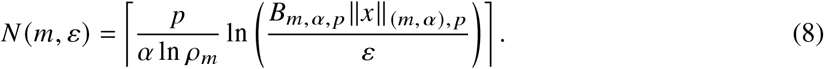

In particular, if *B*_*m,α, p*_ ∥*x*∥_(*m,α*), *p*_ ≤ 1, then the normalized choice 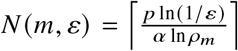 is sufficient. Here, *N* denotes the theoretical truncation dimension and is not assumed to coincide with the number of CNN filters.

### 3.8 Dominant-Root-Based Channel Scaling

Let **F** = (*F*_1_, *F*_2_, … , *F*_7_) = (21, 34, 55, 89, 144, 233, 377) denote the original Fibonacci-Net channel vector. For *m* ≥ 2, define the dominant-root scaling coefficient by

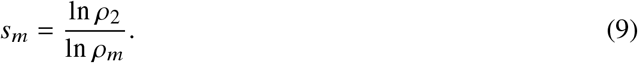

Thus, *s*_2_ = 1 whereas, for *m* > 2, 0 < *s*_*m*_ < 1 and *s*_*m*+1_ < *s*_*m*_

Moreover, 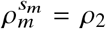 so the exponent *s*_*m*_ maps the dominant growth base *ρ*_*m*_ to the Fibonacci base *ρ*_2_. Accordingly, *s*_*m*_ provides a theoretically motivated architectural scaling rule, while the raw higher-order *m*-Bonacci terms are not used directly as CNN filter counts.

### 3.9 Channel Quantization and Allocation

To obtain hardware-compatible channel dimensions, we define

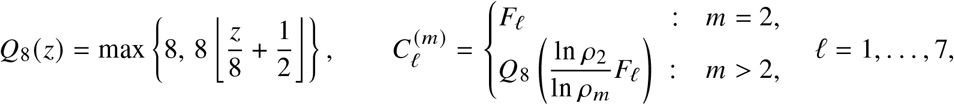

where 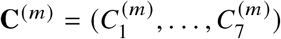. For *m* = 2, … , 6,

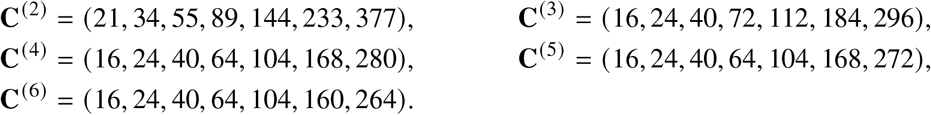

### 3.10 Network Architecture

Let (*f*_1_, *f*_2_, … , *f*_7_) = **C**^(*m*)^. For convolutional blocks 1–5, the standard transformation is defined by

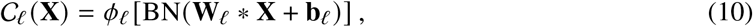

where * denotes convolution, BN batch normalization, and *ϕ*_*l*_ the activation function. The custom pooling operator is

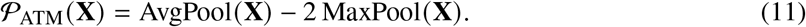

After the second block, the first pooling branch and concatenated feature map are given by

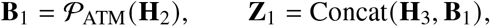

so that *C*(**Z**_1_) = *f*_3_ + *f*_2_.

In addition, a fixed 16-channel skip branch is defined as **S** = *P*_ATM_ [C_*s*_ (**H**_3_)] , where C_*s*_ is a 3 × 3 convolution producing 16 output channels. The second concatenated feature map is therefore

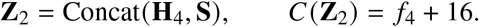

The overall proposed network framework is illustrated in Figure 1

**Figure 1:**
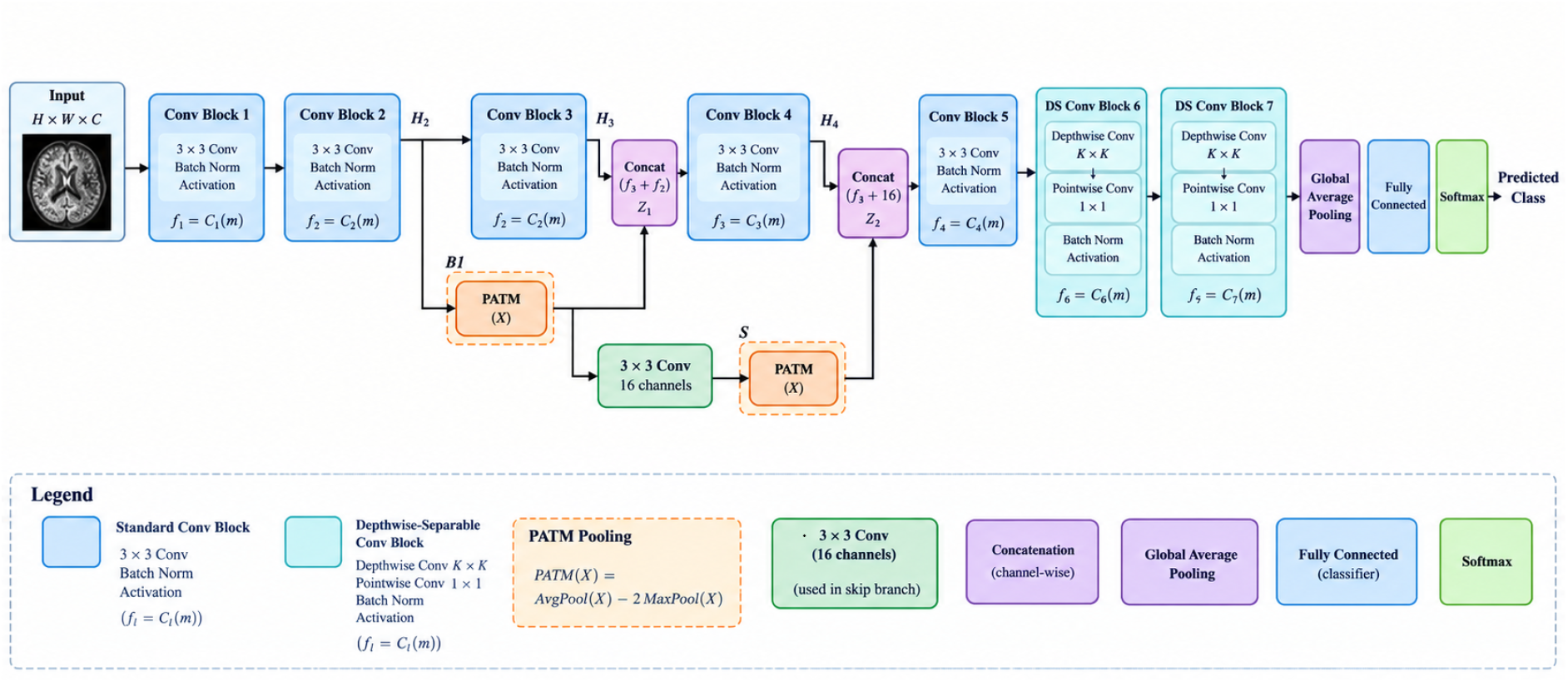
Overview of the proposed methodology.

### 3.11 Depthwise-Separable Convolution

Blocks 6 and 7 employ depthwise-separable convolutions, defined as

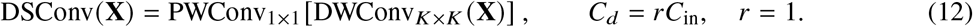

The two blocks, global average pooling, and classification output are given by

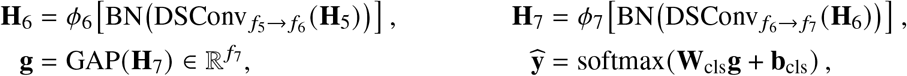

where *N*_*c*_ denotes the number of classes.

### 3.12 Parameter-Count Analysis

For a standard *K* × *K* convolution with bias, the parameter count is

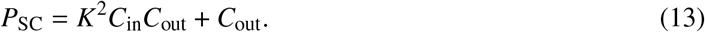

Accordingly, the first five principal convolutions contain

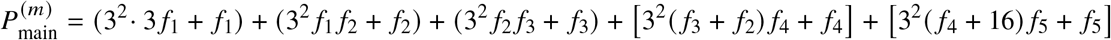

whereas the skip convolution contributes 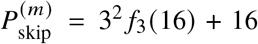. Assuming no separate depthwise bias and one output bias following the pointwise operation, a depthwise-separable convolution has *P*_DS_ = *K*^2^*C*_in_*r* + *rC*_in_*C*_out_ + *C*_out_ parameters.

For, *r* = 1, the parameter counts of blocks 6 and 7 are

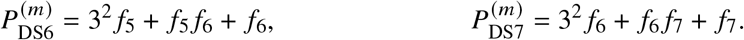

If `use_bias=False` because batch normalization follows each convolution, the final terms *f*_6_ and *f*_7_ are omitted.

The trainable and total stored batch-normalization parameter counts are, respectively,

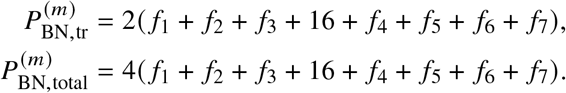

The classifier contributes 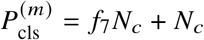, and the total stored parameter count is therefore

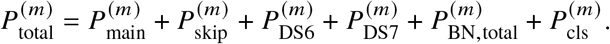

### 3.13 MAC and FLOP Analysis

For a standard convolution producing an *H* × *W* output, the number of multiply-accumulate operations is

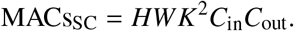

For a depthwise-separable convolution, it is MACs_DS_ = *HW K*^2^*C*_in_*r* + *rC*_in_*C*_out_.

Thus, for *r* = 1, blocks 6 and 7 require

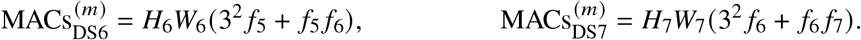

Under the convention that one multiplication and one addition count as two floating-point operations,

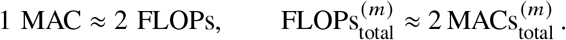

### 3.14 Efficiency of Depthwise-Separable Convolution

Ignoring bias terms, the ratio of depthwise-separable to standard-convolution parameters is

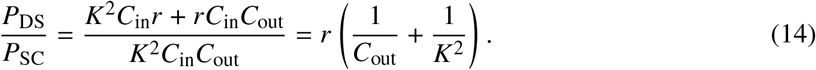

For *r* = 1 and *K* = 3, this reduces to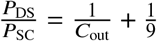.

### 3.15 Complexity Reduction Relative to Fibonacci-Net

The percentage reductions in parameters, MACs, and FLOPs are defined, respectively, as

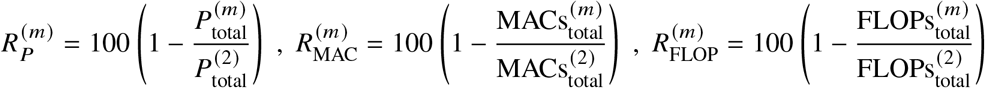

The proposed lightweight-design mechanism can be summarized as

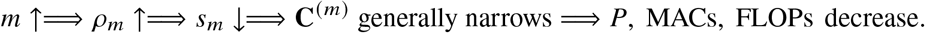

The dominant-root scaling rule determines the order-dependent channel configuration, while depthwise-separable convolution reduces the computational cost of the final convolutional blocks. Preservation of classification performance is an empirical question and is not guaranteed by the mathematical scaling rule alone.

The dominant roots, scaling coefficients, and adopted channel configurations for all evaluated recurrence orders are summarized in Table 1.

**Table 1:**
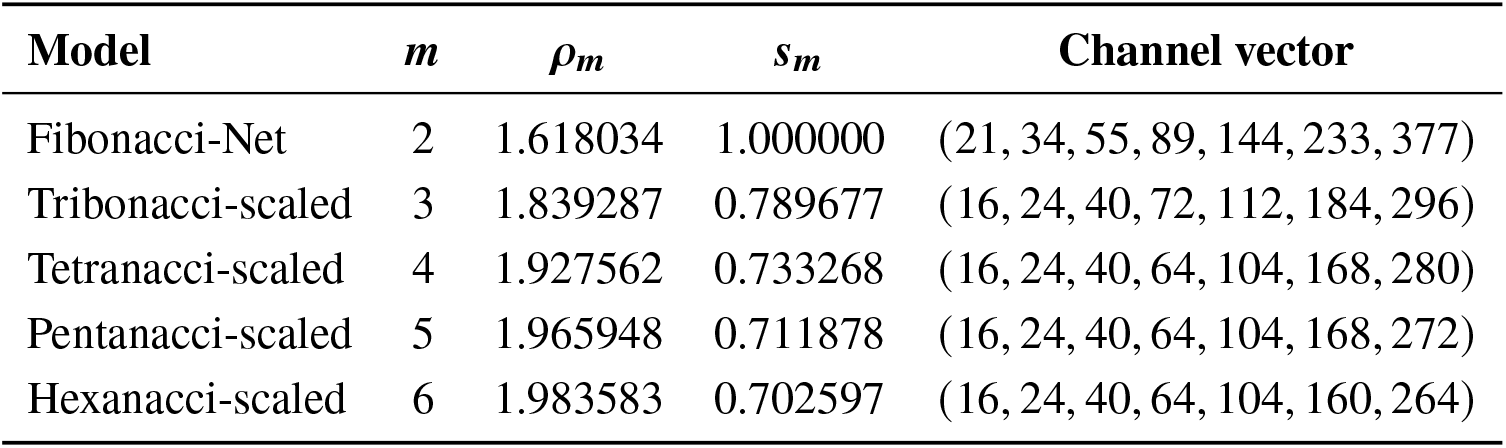
Adopted dominant-root-scaled channel configurations.

| **Model** | ***m*** | ***ρ<sub>m</sub>*** | ***s<sub>m</sub>*** | **Channel vector** |
|---|---|---|---|---|
| Fibonacci-Net | 2 | 1.618034 | 1.000000 | (21, 34, 55, 89, 144, 233, 377) |
| Tribonacci-scaled | 3 | 1.839287 | 0.789677 | (16, 24, 40, 72, 112, 184, 296) |
| Tetranacci-scaled | 4 | 1.927562 | 0.733268 | (16, 24, 40, 64, 104, 168, 280) |
| Pentanacci-scaled | 5 | 1.965948 | 0.711878 | (16, 24, 40, 64, 104, 168, 272) |
| Hexanacci-scaled | 6 | 1.983583 | 0.702597 | (16, 24, 40, 64, 104, 160, 264) |

## 4 Experimental Evaluation of Scaled *m*-BonacciNet

This section evaluates the performance of the proposed Scaled *m*-BonacciNet. It first describes the datasets, preprocessing procedures, experimental setup, and evaluation metrics. The resulting performance is then analysed and compared with that of existing methods across the selected datasets. Additional experiments are conducted to examine the contribution of the model’s key components and its behaviour under different experimental conditions.

### 4.1 Dataset and Implementation Details

The proposed Scaled *m*-BonacciNet was evaluated using two medical image datasets. The first is a publicly available brain MRI dataset [30] comprising 4,479 images from multiple MRI modalities, including T1, T1C+, and T2. By considering each imaging modality separately, the dataset contains 44 highly imbalanced classes, with class sizes ranging from 17 to 233 images. The second dataset is a locally curated ultrasound (US) kidney dataset containing stage-wise chronic kidney disease (CKD) and normal kidney images. The images were collected at Shivam Diagnostic Centre, Jalgaon, Maharashtra, India, with institutional approval and written patient consent. Two radiologists, each with 15 years of experience, supervised the collection, classification, and labelling of the images.

All experiments were implemented using the Keras Sequential API and executed on NVIDIA Tesla P100 GPU. The images were resized to 224 × 224 pixels and divided into training, validation, and testing sets using a stratified ratio of 60:20:20. The models were trained using the Adam optimizer with a batch size of 8 for 50 epochs. The learning rate was initially fixed at 1 × 10^−4^ for the first 13 epochs and subsequently decayed by a factor of 0.9 until the final epoch. No early stopping, data augmentation, or additional image-processing method was applied.

### 4.2 Results and Discussion

Figure 2(a) summarises the accuracy–efficiency characteristics of the scaled *m*-BonacciNet variants across the 4-class, 15-class, and 44-class Brain MRI classification settings. Fibonacci-Net (*m* = 2) was used as the reference model; therefore, its cells are marked as “Ref.” Positive values in Figure 2(a) indicate improvements in test accuracy relative to Fibonacci-Net, whereas negative values indicate reductions. For the 4-class dataset, the scaled variants exhibited accuracy reductions ranging from 0.77 to 2.11 percentage points. Tribonacci-Net (*m* = 3) showed the smallest reduction of 0.77 percentage points, followed by Pentanacci-Net (*m* = 5) with a reduction of 1.15 percentage points, whereas Tetranacci-Net (*m* = 4) and Hexanacci-Net (*m* = 6) each showed a reduction of 2.11 percentage points. On the 15-class dataset, Tribonacci-Net achieved the largest accuracy improvement of 0.67 percentage points, whereas the remaining variants showed reductions of 0.22–0.44 percentage points. On the more challenging 44-class dataset, Pentanacci-Net improved accuracy by 0.34 percentage points, whereas the other variants showed reductions ranging from 0.11 to 0.67 percentage points.

**Figure 2:**
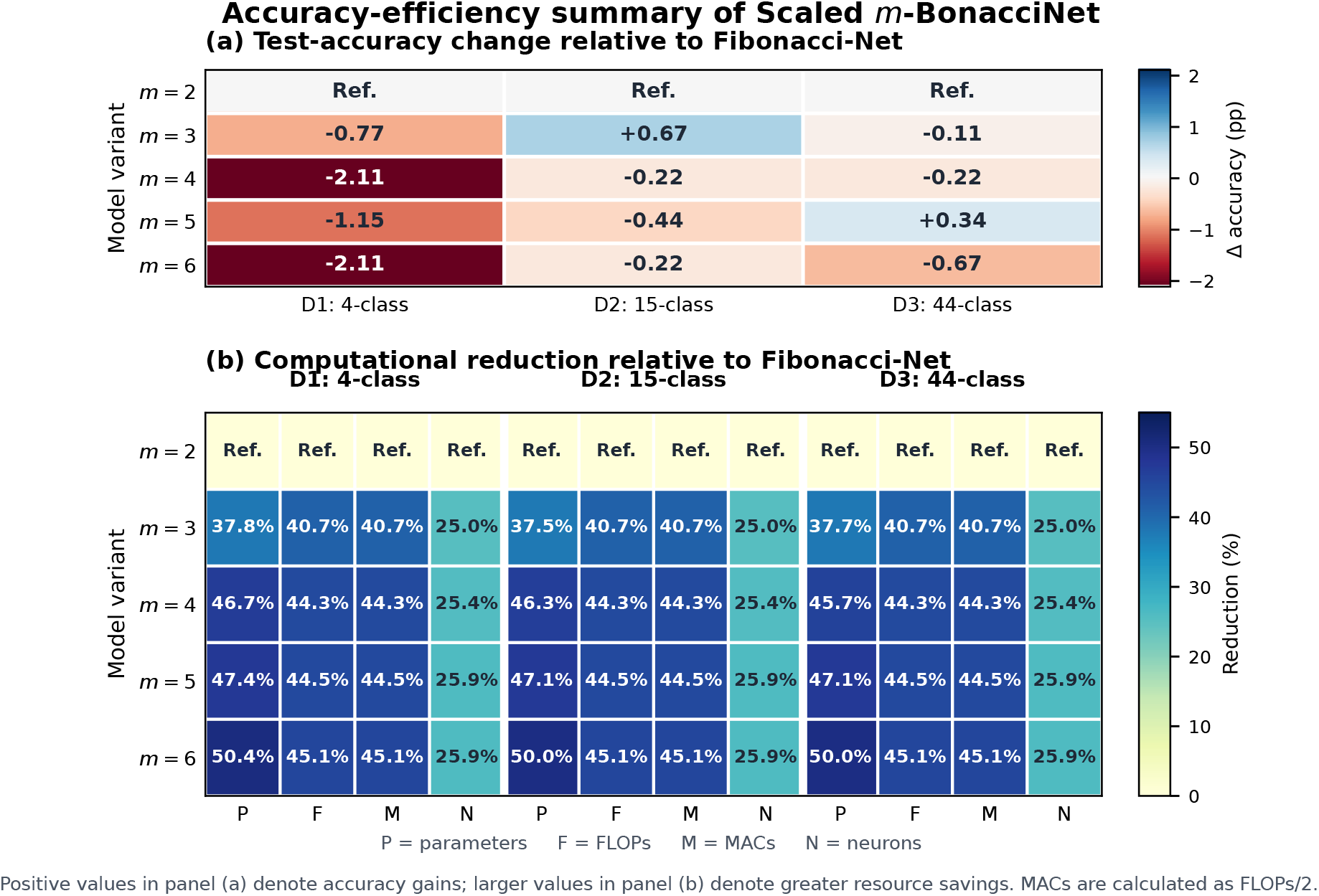
Accuracy–efficiency comparison of the scaled m-BonacciNet variants across three Brain MRI classification settings: D1 (4-class), D2 (15-class), and D3 (44-class).

As shown in Figure 2(b), increasing the recurrence order substantially reduced the computational requirements relative to Fibonacci-Net. Across the three datasets, the scaled variants reduced the number of parameters by approximately 37.5–50.4%, while FLOPs and MACs were reduced by approximately 40.7–45.1%. The reduction in the number of neurons ranged from 25.0% to 25.9% across the three datasets. In general, the computational reductions tended to increase with *m*, with Hexanacci-Net (*m* = 6) providing the greatest overall computational savings. However, Hexanacci-Net produced accuracy reductions of 2.11, 0.22, and 0.67 percentage points on the 4-class, 15-class, and 44-class datasets, respectively.

Overall, the results demonstrate a clear trade-off between predictive accuracy and computational efficiency. Tribonacci-Net provided the most favorable balance on the 15-class dataset, improving accuracy by 0.67 percentage points while reducing parameters by approximately 37.5% and FLOPs and MACs by approximately 40.7%. On the 44-class dataset, Pentanacci-Net offered the most favorable balance, improving accuracy by 0.34 percentage points while reducing parameters by approximately 47.1%, FLOPs and MACs by approximately 44.5%, and the number of neurons by 25.9%. Although none of the scaled variants improved accuracy on the 4-class dataset, they achieved substantial computational reductions. Among these variants, Tribonacci-Net (*m* = 3) produced the smallest accuracy reduction of 0.77 percentage points. These findings indicate that the preferred recurrence order depends on the required balance between classification performance and computational cost.

Table 2 compares the scaled *m*-BonacciNet variants across the 4-, 15-, and 44-class Brain MRI datasets. For the 4-class dataset, Fibonacci-Net achieved the highest test accuracy (94.65%), while the higher-order variants substantially reduced computational complexity with modest reductions in accuracy. For the 15-class dataset, Tribonacci-Net provided the most favorable accuracy-efficiency balance, achieving 96.09% accuracy with 0.85 million parameters and 365.69 million FLOPs. For the 44-class dataset, Pentanacci-Net achieved the highest accuracy (95.87%) using 0.73 million parameters and 342.47 million FLOPs. Across all three datasets, Hexanacci-Net consistently required the fewest parameters and FLOPs, although this computational advantage was accompanied by some reduction in predictive performance. Across the three benchmark datasets, the intermediate-order variants, particularly Tribonacci-Net and Pentanacci-Net, generally provided the most favorable balance between predictive performance and computational efficiency, consistent with the relative trends shown in Figure 2. To further examine the generalizability of the proposed *m*-BonacciNet framework in a clinically distinct imaging setting, all model variants were evaluated on the locally curated CKD ultrasound dataset described in Section 4.1. Table 3 summarizes their classification performance and computational requirements, while Figure 4(a) and Figure 4(b) present the corresponding accuracy efficiency trade-offs relative to Fibonacci-Net.

**Table 2:**
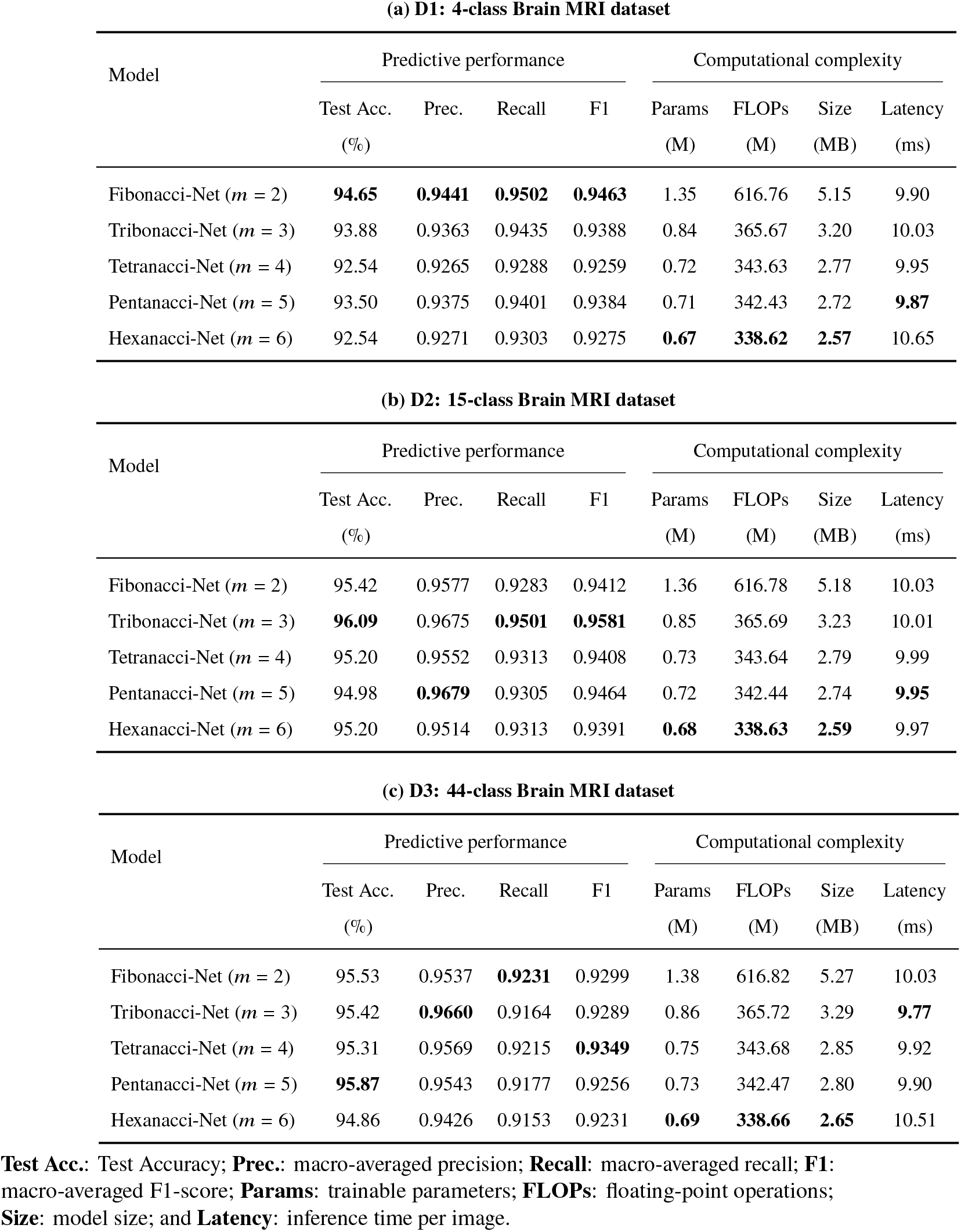
Predictive performance and computational complexity of the scaled *m*-BonacciNet variants across three Brain MRI datasets.

| **(a) D1: 4-class Brain MRI dataset** |  |  |  |  |  |  |  |  |
|---|---|---|---|---|---|---|---|---|
| Model | Predictive performance |  |  |  | Computational complexity |  |  |  |
|  | Test Acc.<br>(%) | Prec. | Recall | F1 | Params<br>(M) | FLOPs<br>(M) | Size<br>(MB) | Latency<br>(ms) |
| Fibonacci-Net (*m* = 2) | **94.65** | **0.9441** | **0.9502** | **0.9463** | 1.35 | 616.76 | 5.15 | 9.90 |
| Tribonacci-Net (*m* = 3) | 93.88 | 0.9363 | 0.9435 | 0.9388 | 0.84 | 365.67 | 3.20 | 10.03 |
| Tetranacci-Net (*m* = 4) | 92.54 | 0.9265 | 0.9288 | 0.9259 | 0.72 | 343.63 | 2.77 | 9.95 |
| Pentanacci-Net (*m* = 5) | 93.50 | 0.9375 | 0.9401 | 0.9384 | 0.71 | 342.43 | 2.72 | **9.87** |
| Hexanacci-Net (*m* = 6) | 92.54 | 0.9271 | 0.9303 | 0.9275 | **0.67** | **338.62** | **2.57** | 10.65 |
| **(b) D2: 15-class Brain MRI dataset** |  |  |  |  |  |  |  |  |
| Model | Predictive performance |  |  |  | Computational complexity |  |  |  |
|  | Test Acc.<br>(%) | Prec. | Recall | F1 | Params<br>(M) | FLOPs<br>(M) | Size<br>(MB) | Latency<br>(ms) |
| Fibonacci-Net (*m* = 2) | 95.42 | 0.9577 | 0.9283 | 0.9412 | 1.36 | 616.78 | 5.18 | 10.03 |
| Tribonacci-Net (*m* = 3) | **96.09** | 0.9675 | **0.9501** | **0.9581** | 0.85 | 365.69 | 3.23 | 10.01 |
| Tetranacci-Net (*m* = 4) | 95.20 | 0.9552 | 0.9313 | 0.9408 | 0.73 | 343.64 | 2.79 | 9.99 |
| Pentanacci-Net (*m* = 5) | 94.98 | **0.9679** | 0.9305 | 0.9464 | 0.72 | 342.44 | 2.74 | **9.95** |
| Hexanacci-Net (*m* = 6) | 95.20 | 0.9514 | 0.9313 | 0.9391 | **0.68** | **338.63** | **2.59** | 9.97 |
| **(c) D3: 44-class Brain MRI dataset** |  |  |  |  |  |  |  |  |
| Model | Predictive performance |  |  |  | Computational complexity |  |  |  |
|  | Test Acc.<br>(%) | Prec. | Recall | F1 | Params<br>(M) | FLOPs<br>(M) | Size<br>(MB) | Latency<br>(ms) |
| Fibonacci-Net (*m* = 2) | 95.53 | 0.9537 | **0.9231** | 0.9299 | 1.38 | 616.82 | 5.27 | 10.03 |
| Tribonacci-Net (*m* = 3) | 95.42 | **0.9660** | 0.9164 | 0.9289 | 0.86 | 365.72 | 3.29 | **9.77** |
| Tetranacci-Net (*m* = 4) | 95.31 | 0.9569 | 0.9215 | **0.9349** | 0.75 | 343.68 | 2.85 | 9.92 |
| Pentanacci-Net (*m* = 5) | **95.87** | 0.9543 | 0.9177 | 0.9256 | 0.73 | 342.47 | 2.80 | 9.90 |
| Hexanacci-Net (*m* = 6) | 94.86 | 0.9426 | 0.9153 | 0.9231 | **0.69** | **338.66** | **2.65** | 10.51 |

**Test Acc.**: Test Accuracy; **Prec.**: macro-averaged precision; **Recall**: macro-averaged recall; **F1**: macro-averaged F1-score; **Params**: trainable parameters; **FLOPs**: floating-point operations; **Size**: model size; and **Latency**: inference time per image.

**Table 3:** Predictive performance and computational complexity of the scaled *m*-BonacciNet variants on the CKD ultrasound dataset.

| Model | Predictive performance |  |  |  | Computational complexity |  |  |  |
|---|---|---|---|---|---|---|---|---|
|  | Test Acc.<br>(%) | Prec. | Recall | F1 | Params<br>(M) | FLOPs<br>(M) | Size<br>(MB) | Latency<br>(ms) |
| Fibonacci-Net (*m* = 2) | 80.00 | 0.8041 | 0.7664 | 0.7744 | 1.35 | 616.76 | 5.16 | 10.73 |
| Tribonacci-Net (*m* = 3) | 75.00 | **0.8055** | 0.7083 | 0.7010 | 0.84 | 365.68 | 3.21 | 10.36 |
| Tetranacci-Net (*m* = 4) | 75.00 | 0.7206 | 0.7069 | 0.6970 | 0.73 | 343.63 | 2.77 | 10.40 |
| Pentanacci-Net (*m* = 5) | 73.75 | 0.6989 | 0.7077 | 0.6914 | 0.71 | **342.43** | 2.72 | 10.46 |
| Hexanacci-Net (*m* = 6) | **83.75** | 0.7942 | **0.7952** | **0.7941** | **0.67** | 338.62 | **2.57** | **10.28** |

**Figure 3:**
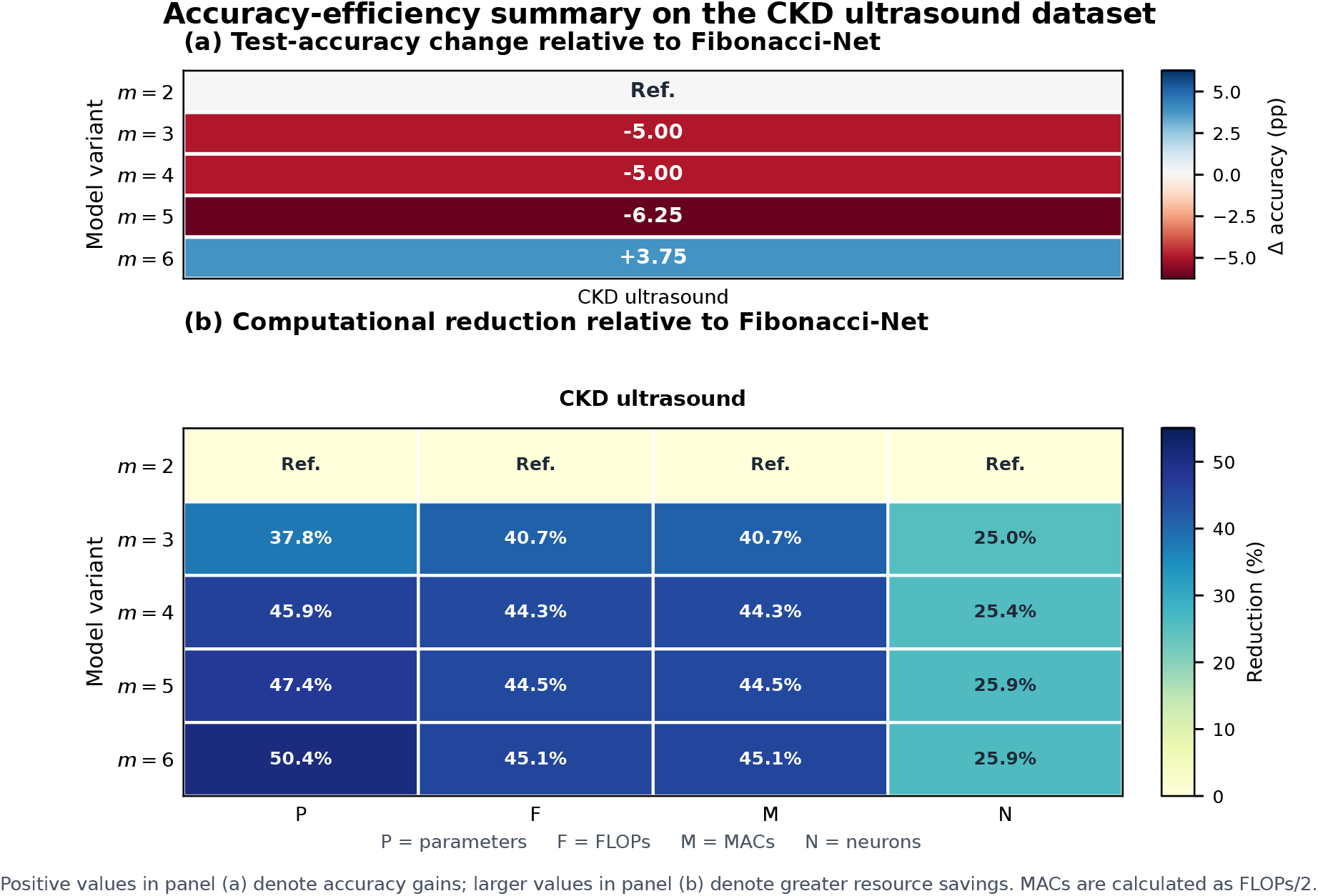
Accuracy efficiency comparison of *m*-BonacciNet variants on the CKD ultrasound dataset: (a) test-accuracy change and (b) computational reductions relative to Fibonacci-Net.

**Figure 4:**
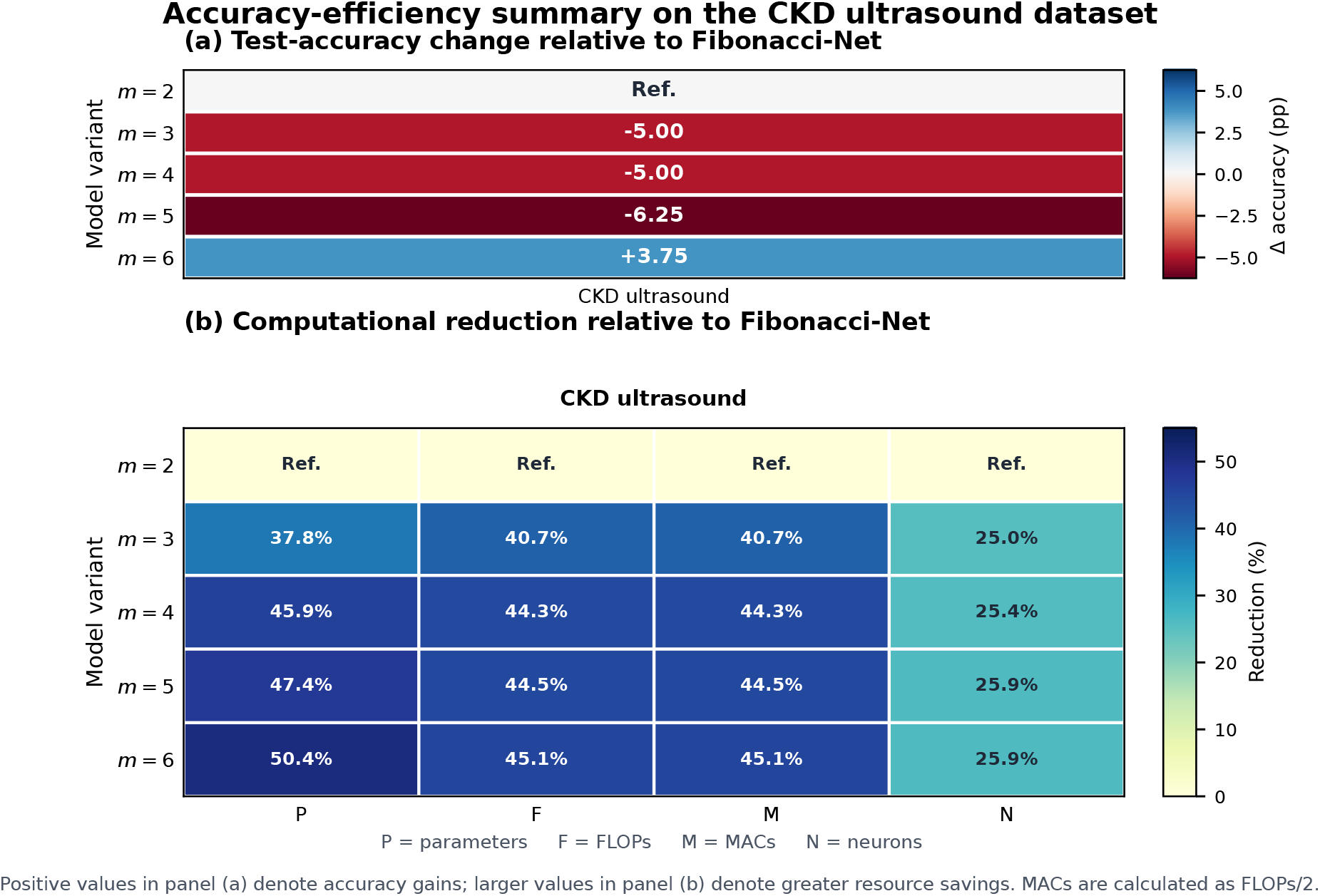
Accuracy efficiency comparison of *m*-BonacciNet variants on the CKD ultrasound dataset: (a) test-accuracy change and (b) computational reductions relative to Fibonacci-Net.

Across the three benchmark datasets, the intermediate-order variants, particularly Tribonacci-Net and Pentanacci-Net, generally provided the most favorable balance between predictive performance and computational efficiency, consistent with the relative trends shown in Figure 2. To further examine the generalizability of the proposed *m*-BonacciNet framework in a clinically distinct imaging setting, all model variants were evaluated on the locally curated CKD ultrasound dataset described in Section 4.1. Table 3 summarizes their classification performance and computational requirements, while Figure 4(a) and Figure 4(b) present the corresponding accuracy efficiency trade-offs relative to Fibonacci-Net.

As shown in Table 3, Hexanacci-Net achieved the highest test accuracy of 83.75%, together with the highest recall and macro-F1 score. It also required the fewest parameters, had the smallest model size, and achieved the lowest latency. Although Tribonacci-Net obtained the highest macro-averaged precision, Hexanacci-Net required the fewest parameters and FLOPs and provided the best overall balance between predictive performance and computational efficiency on the CKD ultrasound dataset.

Unlike the pattern observed across the benchmark datasets, Hexanacci-Net achieved the most favorable overall performance on the CKD ultrasound dataset. It increased test accuracy by 3.75 percentage points relative to Fibonacci-Net while reducing the numbers of parameters, FLOPs, MACs, and neurons by 50.4%, 45.1%, 45.1%, and 25.9%, respectively. In contrast, Tribonacci-Net and Tetranacci-Net each exhibited a 5.00-percentage-point reduction in test accuracy, whereas Pentanacci-Net showed a reduction of 6.25 percentage points, despite their substantial computational savings. These findings indicate that the optimal recurrence order is dataset-dependent: intermediate-order variants offered the strongest trade-offs across the three benchmark datasets, whereas the higher-order Hexanacci-Net was better suited to the CKD ultrasound classification task.

### 4.3 Grad-CAM-Based Model Interpretability

Grad-CAM was applied to representative Brain MRI and CKD ultrasound images to examine the image regions contributing most strongly to the model predictions. This analysis was included because classification metrics alone cannot determine whether the model relies on clinically relevant image content or on irrelevant background features and imaging artefacts.

For the Brain MRI dataset, the activation maps generally emphasized intracranial regions and areas surrounding visible abnormalities (Figure 5a). For the CKD ultrasound dataset, the strongest responses were predominantly distributed within the renal region and around areas showing altered echogenic patterns (Figure 5b).

**Figure 5:**
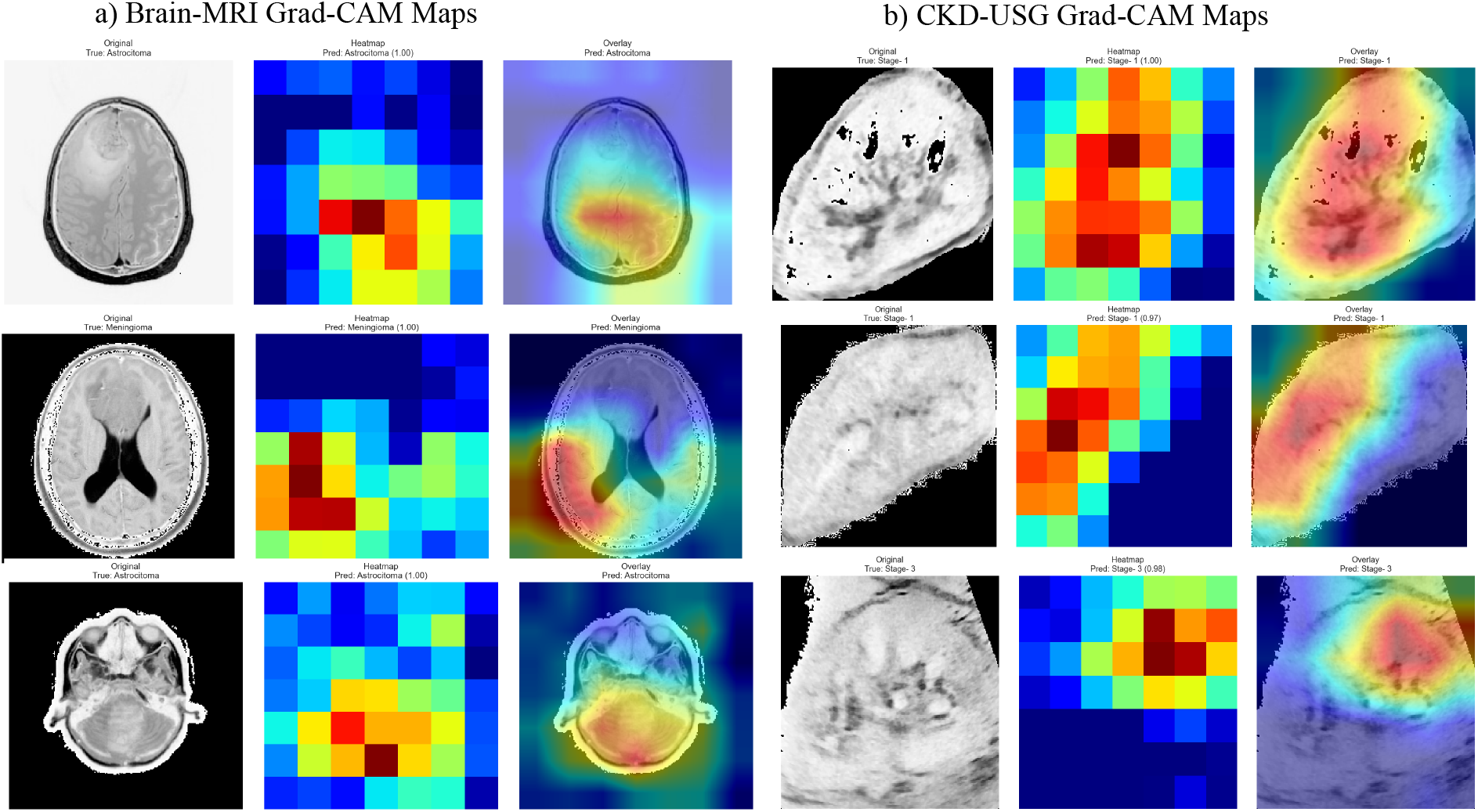
Representative Grad-CAM visualizations for the two imaging datasets: (a) Brain MRI and (b) CKD ultrasound. Each example presents the original image, activation map, and corresponding overlay. Warmer colours indicate image regions contributing more strongly to the model prediction.

Overall, the Grad-CAM results provide qualitative evidence that the predictions were influenced primarily by image regions containing potentially relevant anatomical and pathological information. Nevertheless, these visualizations are explanatory rather than clinically confirmatory and should not be interpreted as lesion-segmentation results or as a substitute for validation by experienced radiologists

## 5 Conclusion

In this study, we introduced the *m*-Bonacci weighted sequence space 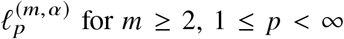 for *m* ≥ 2, 1 ≤ *p* < ∞, and *α* > 0. We established that 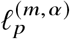 is a Banach coordinate space isometrically isomorphic to *l*_*p*_. Consequently, it possesses an unconditional Schauder basis and, for 1 < *p* < ∞, inherits the reflexivity and uniform convexity of *l*_*p*_. We also established the strict inclusion chain

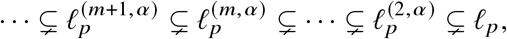

which is governed by the monotonic growth of the dominant *m*-Bonacci roots.

The weighted-space formulation further yields an asymptotic truncation estimate of order 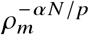 in the unweighted *l*_*p*_ norm. This estimate concerns the theoretical truncation dimension and is not interpreted directly as the number of CNN filters.

Using the dominant root *ρ*_*m*_, we defined the scaling coefficient

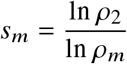

and applied it to the original Fibonacci-Net channel configuration. The resulting adopted channel vectors progressively narrow the network as the recurrence order increases. Depthwise-separable convolution was also employed in the final convolutional blocks to further reduce the parameter and computational requirements.The efficacy of these architectural changes to preserve classification performance while reducing model complexity is evaluated by the experimental results.

## Data Availability

Data is available. Two datasets are used one is publicly available, other one will be availabe on request.

https://www.kaggle.com/datasets/masoudnickparvar/brain-tumor-mri-dataset

## 6 Declarations

## Ethics Approval and Consent to Participate

The permission to use the kidney ultrasound image dataset for academic research was obtained from Shivam Diagnostics, Jalgaon, Maharashtra, India, as documented in a permission letter dated 24 March 2018. The study used the data set for research purposes in accordance with the permission granted by the data provider.

